# Reply to: The chemotherapeutic drug CX-5461 is a potent mutagen in cultured human cells

**DOI:** 10.1101/2025.09.30.25336525

**Authors:** Sehrish Kanwal, Natalie Brajanovski, Jiajun Zhan, Joanna Chan, Richard Rebello, Oliver Hofmann, Sean Grimmond, Luc Furic, Ross D Hannan, Grant McArthur, Gretchen Poortinga, Simon Harrison, Elaine Sanij, Amit Khot, Shahneen Sandhu

**Author notes:** co-senior (Elaine Sanij, Amit Khot, Shahneen Sandhu).

## Abstract

Replying to GCC Koh et al., Nature Genetics (www.nature.com/articles/s41588-023-01602-9) and the comments by Simon Boulton in Nature Genetics (www.nature.com/articles/s41588-023-01611-8).

The drug CX-5461 [Pidnarulex] is an inhibitor of RNA polymerase I and topoisomerase II and a DNA G-quadruplex stabilizer. Koh et al. recently reported that CX-5461 induces extensive, non-selective collateral mutagenesis *in vitro* at magnitudes surpassing known environmental carcinogens, raising concerns about its potential to promote secondary cancers.

Mindful that the report of Koh et al. was exclusively an *in vitro* study, we applied the same ultra-sensitive, error-corrected TwinStrand Duplex Sequencing approach and analyses (methods) to a longitudinal series of clinical specimens from four patients who participated in the Phase I dose-escalation trial of CX-5461 in advanced haematologic malignancies [ACTRN12613001061729]. In contrast to the findings of Koh et al. we found no evidence that clinical administration of CX-5461 significantly increased mutational burden in patient samples, nor did we observe the reported mutational signature.

These results suggest that the mutagenic effects described *in vitro* do not translate to the clinical setting. This is particularly important given that several clinical trials, including a Phase Ib study of CX-5461 in patients with solid tumours [NCT04890613] and a National Cancer Institute sponsored Phase I trial [NCT06606990] are currently enrolling patients. These ongoing and planned clinical efforts highlight both the therapeutic promise of CX-5461 and the importance of evaluating this agent within rigorous and clinically relevant frameworks.

## Response

Replying to GCC Koh *et al*., Nature Genetics (https://www.nature.com/articles/s41588-023-01602-9) and the comments by Simon Boulton in Nature Genetics (https://www.nature.com/articles/s41588-023-01611-8).

The drug CX-5461 (Pidnarulex) is an inhibitor of RNA polymerase I and topoisomerase II and a DNA G-quadruplex stabilizer^1-7^. GCC Koh *et al*. recently reported that CX-5461 induces extensive, non-selective, collateral mutagenesis in cells *in vitro* at magnitudes surpassing known environmental carcinogens, raising concerns of its secondary cancer promoting potential. They reported that potency was so high that a single acute exposure induced widespread mutations in cultured human cells. Given these potential safety concerns, we elected to suspend our Phase I trial of CX-5461 and Talazoparib (NCT05425862) until the findings by Koh *et al*. could be corroborated. The trial investigators ensured that all participants and treating clinicians were fully informed of the publication’s findings and were provided with appropriate clinical context to support informed decision-making.

Mindful that the report of Koh *et al*. was exclusively an *in vitro* study, we sought to determine the clinical impact of clinical grade CX-5461 exposure. We undertook the same ultra-sensitive, error-corrected TwinStrand Duplex Sequencing^8,9^ approach and analyses (methods) used by Koh *et al*. on a longitudinal series of clinical specimens collected from 4 patients who participated in the Phase I dose-escalation trial of CX-5461 in advanced haematologic malignancies^10^ (ACTRN12613001061729). The dosage range (50-170mg/m^2^) and timepoints of drug exposure included 4h, 24h post Cycle 1 Day 1 and end of treatment (EOT) samples at plasma drug concentrations higher than those used *in vitro* in Koh *et al*. The samples were selected on the basis of drug exposure and the availability of multiple matched normal (skin and bone marrow (BM) aspirates) and tumour tissue and peripheral blood mononucleocytes collected at baseline and post 3-12 weeks at EOT (Supplementary Table 1).

Contrary to the findings of Koh *et al*., using an identical sequencing platform (DuplexSeq), we found no evidence of clinical CX-5461 administration significantly elevating the mutational burdens in patient samples (Figure 1a, Supplementary Table 2). Single base substitutions (SBS) were the most common, with baseline samples prior to exposure to CX-5461 having a slightly higher SBS burden overall than EOT samples (Figure 1a, Supplementary Table 2), DBS analysis of paired pre- and post-treatment samples shows that CX-5461 does not measurably increase DBS burden (Figure 2a, Supplementary Table 2 and 3). All patient samples were higher than the internal TwinStrand control (176 SBS mutations). In most cases DBS and INDEL burdens did not exceed the internal control and were rare (<20 DBS per sample), with the exception of skin biopsies expectedly displaying UV-induced thymine dimer damage. Collectively, these results indicate that therapeutic dosing of CX-5461 does not induce a measurable burden in patients, with all observed variations falling within the range of normal biological fluctuation and technical variability.

**Figure 1.**
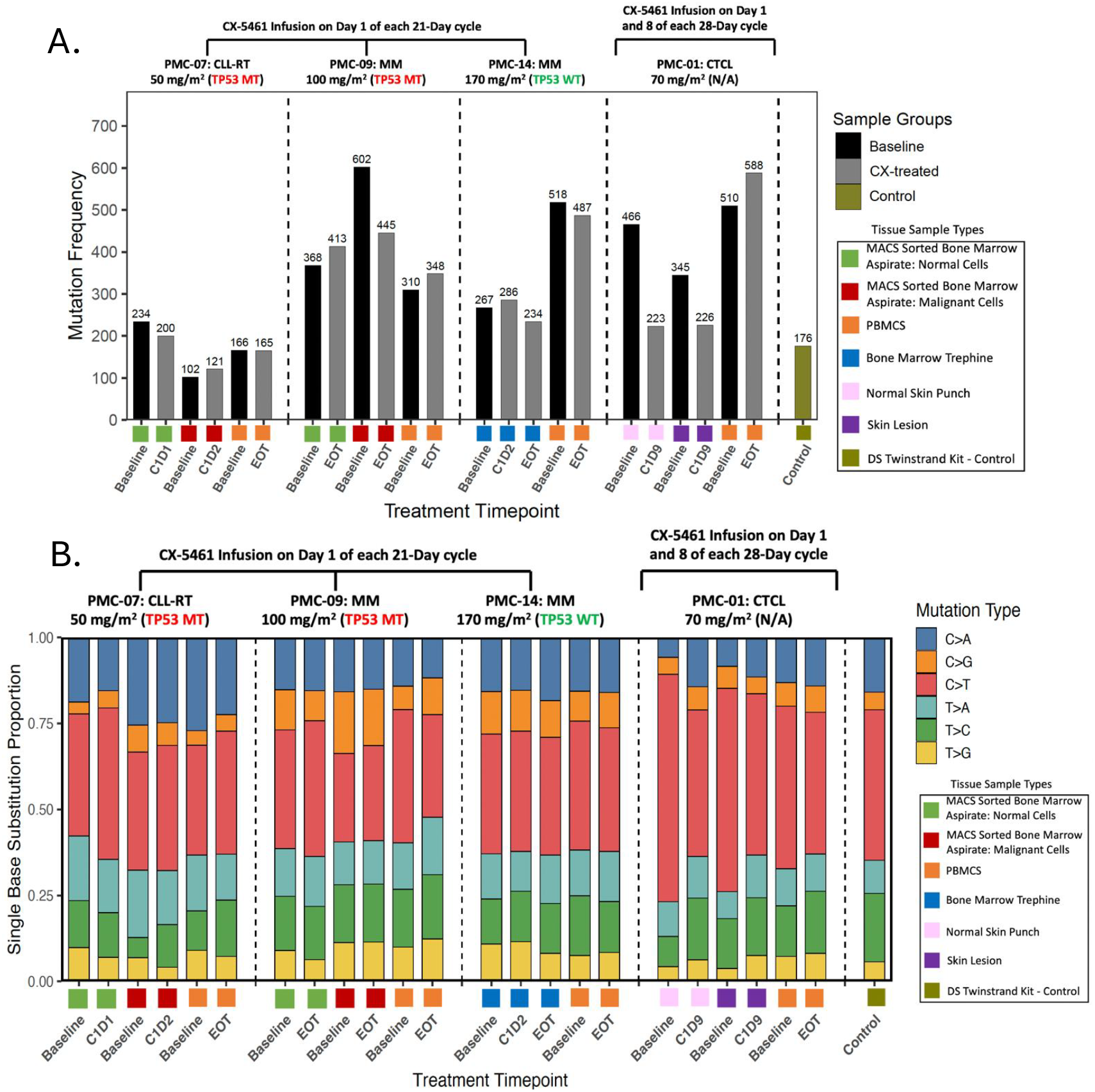
Somatic single-nucleotide variants frequency and proportion across tissue types and treatment timepoints in patients receiving CX-5461. Duplex sequencing was used to quantify single-nucleotide variants (SNVs) across four patients treated with CX-5461 under different dosing regimens. Patients received CX-5461 either on Day 1 of each 21-day cycle (PMC-07: CLL-RT, 50 mg/m^2^, TP53 mutant [Arg248Trp]; PMC-09: MM, 100 mg/m^2^, TP53 mutant [Cys238Tyr]; PMC-14: MM, 170 mg/m^2^, TP53 wild-type), or on Day 1 and Day 8 of each 28-day cycle (PMC-01: CTCL, 70 mg/m^2^, TP53 status not available). **(A.)** Bars represent the absolute number of SNVs detected per sample, grouped by patient and timepoint: Baseline, on-treatment (Cycle 1 Day 1, 2 and 9), and End of Treatment (EOT).). **(B.)** Stacked bar plots show the proportion of six SBS classes (C>A, C>G, C>T, T>A, T>C, T>G) per sample. Bars are grouped by patient and arranged chronologically: Baseline, on-treatment (Cycle 1 Day 1, 2, or 9), and End of Treatment (EOT). Color-coded squares beneath each bar indicate the tissue type sequenced, including MACS-sorted bone marrow (tumor and normal fractions), PBMCs, bone marrow trephines, and paired skin biopsies (lesional and normal). A Duplex Sequencing TwinStrand kit was included as control.

**Figure 2.**
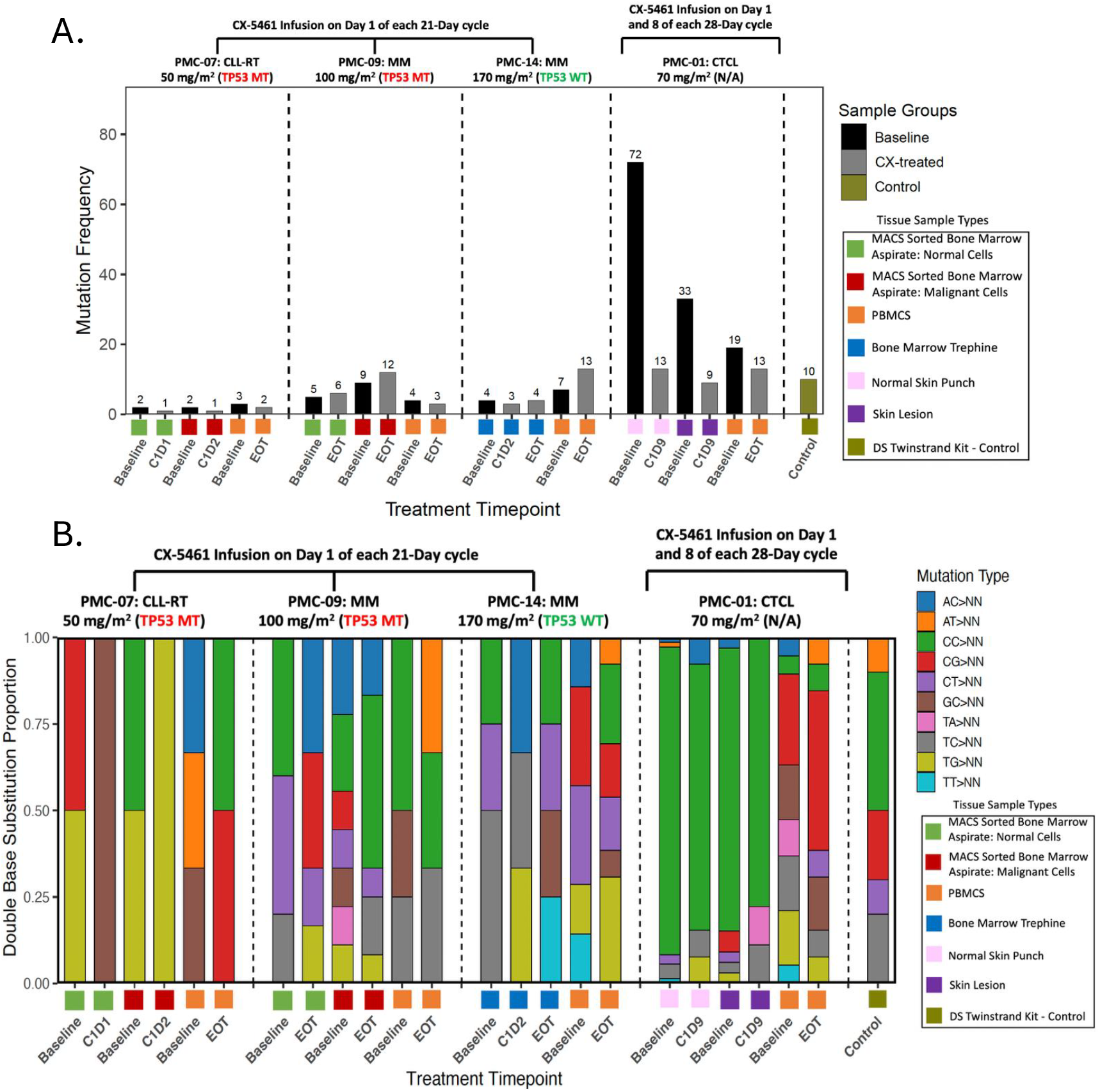
Somatic double base substitution frequency and proportion across tissue types and treatment timepoints in patients receiving CX-5461. Duplex sequencing was used to quantify doublet-base substitutions (DBSs) across four patients treated with CX-5461 under different dosing regimens. Patients received CX-5461 either on Day 1 of each 21-day cycle (PMC-07: CLL-RT, 50 mg/m^2^, TP53 mutant [Arg248Trp]; PMC-09: MM, 100 mg/m^2^, TP53 mutant [Cys238Tyr]; PMC-14: MM, 170 mg/m^2^, TP53 wild-type), or on Day 1 and Day 8 of each 28-day cycle (PMC-01: CTCL, 70 mg/m^2^, TP53 status not available). **(A.)** Bars represent the absolute number of DBSs detected per sample, grouped by patient and timepoint: Baseline, on-treatment (Cycle 1 Day 1, 2 and 9), and End of Treatment (EOT).). **(B.)** Stacked bar plots show the proportional contribution of DBS to the total DBS burden within each sample. Ten mutation types (e.g., AC>NN, CT>NN, TT>NN) are color-coded as indicated in the legend. Bars are grouped by patient and arranged chronologically: Baseline, on-treatment (Cycle 1 Day 1, 2, or 9), and End of Treatment (EOT). Color-coded squares beneath each bar indicate the tissue type sequenced, including MACS-sorted bone marrow (tumor and normal fractions), PBMCs, bone marrow trephines, and paired skin biopsies (lesional and normal). A Duplex Sequencing TwinStrand kit was included as control.

**Figure 3.**
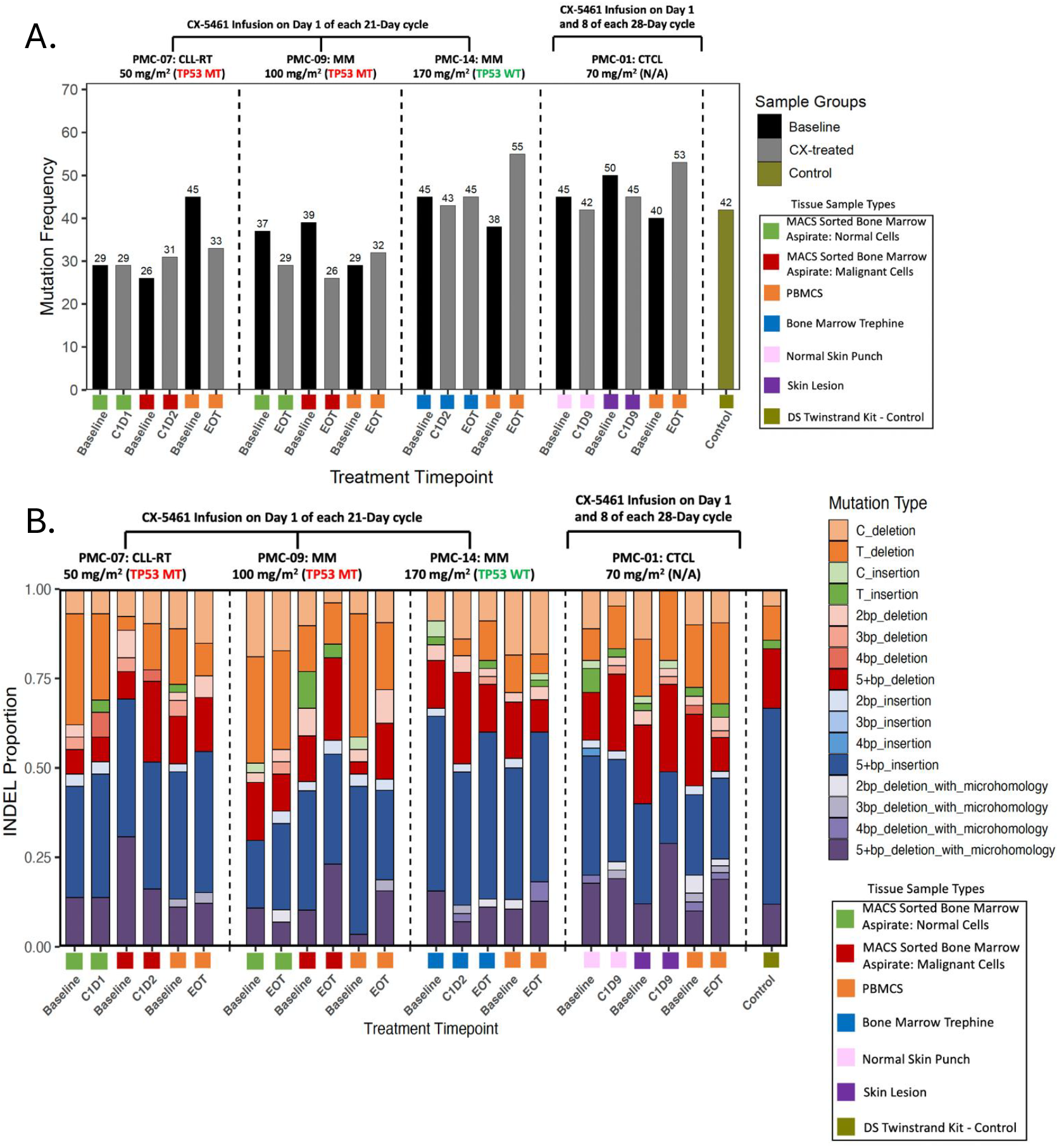
Somatic small insertions and deletions frequency and proportion across tissue types and treatment timepoints in patients receiving CX-5461. Duplex sequencing was used to quantify insertion and deletion (Indel) across four patients treated with CX-5461 under different dosing regimens. Patients received CX-5461 either on Day 1 of each 21-day cycle (PMC-07: CLL-RT, 50 mg/m^2^, TP53 mutant [Arg248Trp]; PMC-09: MM, 100 mg/m^2^, TP53 mutant [Cys238Tyr]; PMC-14: MM, 170 mg/m^2^, TP53 wild-type), or on Day 1 and Day 8 of each 28-day cycle (PMC-01: CTCL, 70 mg/m^2^, TP53 status not available). **(A.)** Bars represent the absolute number of Indels detected per sample, grouped by patient and timepoint: Baseline, on-treatment (Cycle 1 Day 1, 2 and 9), and End of Treatment (EOT).). **(B.)** Stacked bar plots show the proportional composition of Indel classes in each sample. Indels were grouped by type (e.g., C/T base deletions, short/long insertions or deletions) and further annotated for the presence of microhomology (e.g., 2–5+ bp deletions with microhomology), as indicated in the legend. Bars are grouped by patient and arranged chronologically: Baseline, on-treatment (Cycle 1 Day 1, 2, or 9), and End of Treatment (EOT). Color-coded squares beneath each bar indicate the tissue type sequenced, including MACS-sorted bone marrow (tumor and normal fractions), PBMCs, bone marrow trephines, and paired skin biopsies (lesional and normal). A Duplex Sequencing TwinStrand kit was included as control.

Next, we looked for evidence of the SBS, DBS and INDEL mutation profiles Koh *et al*. reported as being specifically induced by CX-5461 exposure. We found no evidence of the reported SBS-mutational signature, which is characterized predominantly by T>A and T>C simple base substitutions with enrichment in ATA and ATG trinucleotide contexts (Supplementary Table 2). Furthermore, the hallmark AT > CA/GA/TA and TG > AT/CT/GT substitutions were not observed in any patient sample across tissue types or dosing levels. Similarly, the reported hallmark CX-5461-induced INDEL signature (InD-CX-5461) consisting of 1 bp T deletions, 2-4 bp duplications, and microhomology-mediated deletions, was not enriched in any post-treatment sample. The low level infrequent INDEL events detected were confined to isolated samples and consistent with stochastic variation rather than treatment-induced enrichment. In summary, no evidence was found to support CX-5461 is a potent non-selective mutagen *in vivo*. None of the previously documented signatures could be reproduced in our clinical cohort, highlighting a discrepancy between the *in vitro* studies from Koh *et al*. and clinical samples.

Scientific claims with direct implications on clinical care must be held to rigorous standards and, at the very least, approximate physiological relevance, particularly when they may cause alarm among vulnerable patients, many of whom are receiving CX-5461 as a potential disease altering anticancer therapy. It is concerning to us that the Koh *et al*. manuscript did not adequately address the study’s significant limitations. These include the fact that there was no evidence presented to demonstrate that the drug concentrations and exposure timelines used in *in vitro* cell lines are relevant to therapeutic exposure in patients. Koh and colleagues did not use clinical grade CX-5461 and the presence of potential contaminants could account for the mutagenic signature they observed. Moreover, the cell lines used in the study, wild-type, *BRCA1*-deficient or *BRCA2*-deficient human hTERT-RPE1 cells and haploid HAP1 cells, do not adequately represent physiological human tissue.

The findings we present here are particularly significant given that two clinical trials of CX-5461 are currently enrolling patients. In a Phase Ib study involving heavily pre-treated patients with advanced solid tumours including those with germline *BRCA1/2* mutations, CX-5461 achieved a clinical benefit rate of 40%^7^. A Phase Ib study of CX-5461 in patients with homologous recombination (HR) deficient solid tumours (NCT04890613) and a National Cancer Institute–sponsored Phase I trial in metastatic solid tumours (NCT06606990) are enrolling patients. The latter study aims to evaluate the safety profile and define the optimal dose of CX-5461. These ongoing and planned clinical efforts underscore the therapeutic promise of CX-5461 and this class of drugs. We hope that our examination of clinical material provides a critical counterbalance to the concerns raised by Koh *et al*., reinforcing the importance of evaluating this agent within rigorous and clinically relevant frameworks.

We unreservedly support the need for greater testing of therapeutic agents prior to use in humans, especially with the recent advances in molecular profiling of mutagenicity. What we would like to emphasise is the need to introduce greater rigour and clinical relevance to these approaches. The Koh *et al*. study (and accompanying commentary) did not adequately address the critical differences between research-based mutagenicity assessment and regulatory safety evaluations of this drug, where CX-5461 was deemed non-mutagenic in GLP (Good Laboratory Practice)-compliant studies. Such testing was performed prior to ethical approval to undertake a human clinical trial with a novel agent. Crucially, evidence demonstrating CX-5461’s low non-selective mutagenic potential in model systems was previously reported in a C. *elegans* study looking at a series of drugs and genetic backgrounds^11^. Finally, it is important to note that in clinical practice, it is both established and accepted that unfortunately many highly active anticancer drugs can increase the risk for developing secondary cancers (anthracyclines, etoposide and the immunomodulatory drug lenalidomide are notable examples)^12-14^. Ultimately, patients are educated and consented about these risks and clinicians make informed decisions by weighing risks against potential therapeutic benefits.

## Author Contributions

S.K. and N.B. performed sample processing and data preparation; S.S., S.G. and A.K. supervised the work. Data interpretation and write-up provided by S.K., N.B., S.S., E.S and S.G. with input from all the other authors, who had the opportunity to edit the manuscript and approved of the submitted version

## Supplementary Information

### Materials and Methods

#### Duplex Sample Set

Clinical sample sets from four patients with advanced haematological malignancies were provided for error-corrected TwinStrand duplex sequencing to investigate the mutation frequency and mutation spectrum in matched normal and malignant tissue, before and after CX-5461 treatment. These samples were obtained from patients enrolled in the first-in-human Phase I dose-escalation study conducted at the Peter MacCallum Cancer Centre (Melbourne, Australia) (ACTRN12613001061729). The treatment protocol initially consisted of CX-5461 administration via a one-hour intravenous infusion on day 1 of each 21-day cycle (patients PMC-07, PMC-09, PMC-14)^10^. Following a protocol amendment, the regimen was modified to administration on Days 1 and 8 of a 28-day cycle (patient PMC-01). The patient cohort comprised two cases of Multiple Myeloma (PMC-09 and PMC-14), one case of Chronic Lymphocytic Leukaemia with Richter’s Transformation (PMC-07), and one case of cutaneous T-cell Lymphoma (PMC-01).

#### DNA extraction and quality control

Genomic DNA was isolated from MACS sorted leukocytes, PBMC’s or snap frozen tissue using the DNeasy Blood & Tissue Kit (Qiagen, Cat# 69504). Following manufacturer’s instructions DNA was eluted in low EDTA TE buffer (10 mM Tris-HCl.5 mM EDTA, pH 9.0) and the concentration quantified using the Qubit™ fluorometer instrument with dsDNA High Sensitivity Assay reagents (Q32854; Thermo Scientific).

#### Duplex sequencing library preparation

Isolated DNA was shipped on dry ice to Inotive-RTP (Inotiv, Inc. Research Triangle Park, North Carolina, US) for library preparation and Duplex sequencing. Genomic DNA integrity was firstly assessed by electrophoresis on a 0.8% agarose gel stained with ethidium bromide (0.15 μg/mL) and DNA purity examined using the microvolume spectrophotometer. Samples whose 260/230 absorbance ratio fell below 1.7, had undergone a clean-up step using CleanNGS SPRI Beads (Bulldog Bio, Portsmouth, NH, USA) according to Inotiv RTP SOP. Error corrected duplex sequencing was then performed using The TwinStrand Duplex Sequencing Mutagenesis Panel (Human-50), v2.0 kit (TwinStrand Bioscience, Seattle, WA, USA). The kit comprises of 20 target genomic loci, each 2.4 kb in size, that are distributed across 20 autosomal chromosomes (except chromosomes 3 and 5) and include both genic and intergenic sequences that are representative of the complete genome with regard to % GC-content. Briefly, 650ng of DNA was ultrasonically sheared to a median size of ∼300 bp, end-repaired, A-tailed, and then ligated to DuplexSeq Adapters before undergoing target enrichment using the standard reagents and protocol included with the TwinStrand® DuplexSeq™ Human Mutagenesis kit. Prepared sequencing Libraries were then shipped on dry ice to Psomagen where next generation sequencing was performed using the Illumina NovaSeq X Plus platform (Illumina, San Diego CA, USA). All libraries were pooled together at equimolar concentrations and loaded to yield ∼1–1.25 billion duplex bp per sample.

#### Analysis of Duplex Sequencing Results

FastQ files containing TwinStrand duplex sequencing data were processed and analysed by Inotive-RTP using the TwinStrand DuplexSeq Mutagenesis App (version 4.1.0), hosted on the DNAnexus cloud-based platform. The Mutagenesis App performed error-correction and generated Duplex Consensus alignment and variant calls for both germline and ultra-rare somatic variants. The analysis provided the mutation frequency (MF), and types of base substitutions measured across the 20 bait loci for each sample. MF was calculated as the ratio of mutant duplex bases to total duplex bases for each sample. Only variants with variant allele frequency (VAF) < 1% were considered to be the result of mutagenesis (i.e., mutation), and Single nucleotide variants (SNV) overlapping clonal Multi-nucleotide variants (MNV) (VAF >= 0.01) were not considered to be the result of mutagenesis due to a variant calling artifact generating process. Variant filtering was performed using bcftools (version 1.21) and bedtools (version 2.31.0). These were used for subsequent mutation burden and signature analysis. Comprehensive signature analysis was performed using MutationalPatterns (version 3.12.0) with BSgenome.Hsapiens.UCSC.hg38 (version 3.21) as the reference genome and COSMIC signature v3.2 (March 2021 release) as the reference signature set.

## Data availability

A reporting summary for this article is available as a Supplementary Information File. Data generated and/or analyzed are available from the corresponding authors.

