## Supplementary Tables for "Reply to: The chemotherapeutic drug CX-5461 is a potent mutagen in cultured human cells"

**Supplementary Table 1. Overall Duplex sequencing mutation frequencies in matched tumour and normal samples collected from haematological cancer patients treated with CX-5461**

**CX-5461 Cohort - Part 1** — **IV infusion Day 1 of each 21 day cycle**

| Patient ID | Disease | TP53 Status | Dose | Cycles | Timepoint | Sample Type | Tissue | Duplex Bases | | Mutation Frequency | Fold Change: (post/ pre-treatment) |
|---|---|---|---|---|---|---|---|---|---|---|---|
| | | | | | | | | Total | Mutant | | |
| PMC-07 | Chronic Lymphocytic Leukaemia with Richter's transformation | Mutant (Arg248Trp) | 50mg/m2 | 1 | Baseline | MACS Sorted Bone Marrow Aspirate: Tumour Cells | Tumour | 1,074,473,652 | 132 | 1.229E-07 | |
| | | | | | Cycle 1 Day 2 (+24hr post) | | Tumour | 1,309,309,413 | 154 | 1.176E-07 | 0.96 |
| | | | | | Baseline | MACS Sorted Bone Marrow Aspirate: Normal Cells | Normal | 1,081,638,839 | 264 | 2.441E-07 | |
| | | | | | Cycle 1 Day 1 (+4hr post) | | Normal | 1,214,224,508 | 231 | 1.902E-07 | 0.78 |
| | | | | | Baseline | PBMC | Normal | 1,485,032,709 | 213 | 1.434E-07 | |
| | | | | | End of Treatment | PBMC | Normal | 1,689,093,455 | 202 | 1.196E-07 | 0.83 |
| PMC-09 | Multiple Myeloma | Mutant (Cys238Tyr) | 100mg/m2 | 1 | Baseline | MACS Sorted Bone Marrow Aspirate: Tumour Cells | Tumour | 1,322,493,863 | 654 | 4.945E-07 | |
| | | | | | End of Treatment | | Tumour | 1,093,046,179 | 476 | 4.355E-07 | 0.88 |
| | | | | | Baseline | MACS Sorted Bone Marrow Aspirate: Normal Cells | Normal | 1,173,373,725 | 412 | 3.511E-07 | |
| | | | | | End of Treatment | | Normal | 1,447,943,876 | 445 | 3.073E-07 | 0.88 |
| | | | | | Baseline | PBMC | Normal | 1,254,894,805 | 342 | 2.725E-07 | |
| | | | | | End of Treatment | PBMC | Normal | 1,084,163,177 | 392 | 3.616E-07 | 1.33 |
| PMC-14 | Multiple Myeloma | Wildtype | 170mg/m2 | 4 | Baseline | Bone Marrow Trephine | Tumour | 1,317,148,281 | 316 | 2.399E-07 | |
| | | | | | Cycle 1 Day 2 (+24hr post) | | Tumour | 1,238,207,303 | 331 | 2.673E-07 | 1.11 |
| | | | | | End of Treatment | | Tumour | 1,402,659,897 | 284 | 2.025E-07 | 0.84 |
| | | | | | Baseline | PBMC | Normal | 1,676,723,313 | 559 | 3.334E-07 | |
| | | | | | End of Treatment | PBMC | Normal | 1,414,069,220 | 550 | 3.889E-07 | 1.17 |

**CX-5461 Cohort - Part 2** — **IV infusion Day 1 and Day 8 of each 28 day cycle**

| Patient ID | Disease | TP53 Status | Dose | Cycles | Timepoint | Sample Type | Tissue | Duplex Bases | | Mutation Frequency | Fold Change: (post/ pre-treatment) |
|---|---|---|---|---|---|---|---|---|---|---|---|
| | | | | | | | | Total | Mutant | | |
| PMC-01 | Cutaneous T-cell lymphoma | N/A* | 70mg/m2 | 2 | Baseline | Skin Lesion | Tumour | 1,349,753,065 | 429 | 3.178E-07 | |
| | | | | | Cycle 1 Day 9 (+24hr post) | | Tumour | 1,415,430,490 | 283 | 1.999E-07 | 0.63 |
| | | | | | Baseline | Normal Skin Punch | Normal | 1,531,665,094 | 583 | 3.806E-07 | |
| | | | | | Cycle 1 Day 9 (+24hr post) | | Normal | 1,028,577,469 | 279 | 2.712E-07 | 0.71 |
| | | | | | Baseline | PBMC | Normal | 1,273,785,852 | 561 | 4.404E-07 | |
| | | | | | End of Treatment | PBMC | Normal | 1,561,854,818 | 653 | 4.181E-07 | 0.95 |

N/A*: insufficent DNA available to perform All haem Twist assay to determined TP53 status

**Supplementary Table 2. Somatic single-nucleotide variants (SBS) frequency and proportion across tissue types and treatment timepoints in patients receiving CX-5461.**

**CX-5461 Cohort - Part 1** IV infusion Day 1 of each 21 day cycle

| | | | | | | | | SBS Mutation Frequencies | | | | | | SBS Proportions | | | | | | Ratio of SBS Proportions - Post-Treatment:Pre-Treatment | | | | | |
|---|---|---|---|---|---|---|---|---|---|---|---|---|---|---|---|---|---|---|---|---|---|---|---|---|---|
| **Patient** | **Disease** | **TP53 Status** | **Dose** | **Cycles** | **Timepoint** | **Sample ID** | **Tissue** | **C>A** | **C>G** | **C>T** | **T>A** | **T>C** | **T>G** | **C>A** | **C>G** | **C>T** | **T>A** | **T>C** | **T>G** | **C>A** | **C>G** | **C>T** | **T>A** | **T>C** | **T>G** |
| PMC-07 | Chronic Lymphocytic Leukaemia with Richter's transformation | Mutant (Arg248Trp) | 50mg/m2 | 1 | Baseline | MACS Sorted Bone Marrow Aspirate: | Tumour | 2.42E-08 | 7.45E-09 | 3.26E-08 | 1.86E-08 | 5.58E-09 | 6.51E-09 | 2.55E-01 | 7.84E-02 | 3.43E-01 | 1.96E-01 | 5.88E-02 | 6.86E-02 | | | | | | |
| | | | | | Cycle 1 Day 2 (+24hr post) | Tumour Cells | Tumour | 2.29E-08 | 6.11E-09 | 3.36E-08 | 1.45E-08 | 1.15E-08 | 3.82E-09 | 2.48E-01 | 6.61E-02 | 3.64E-01 | 1.57E-01 | 1.24E-01 | 4.13E-02 | 0.97 | 0.84 | 1.06 | 0.80 | 2.11 | 0.60 |
| | | | | | Baseline | MACS Sorted Bone Marrow Aspirate: | Normal | 4.07E-08 | 7.40E-09 | 7.67E-08 | 4.07E-08 | 2.96E-08 | 2.13E-08 | 1.88E-01 | 3.42E-02 | 3.55E-01 | 1.88E-01 | 1.37E-01 | 9.83E-02 | | | | | | |
| | | | | | Cycle 1 Day 1 (+4hr post) | Normal Cells | Normal | 2.55E-08 | 8.24E-09 | 7.25E-08 | 2.55E-08 | 2.14E-08 | 1.15E-08 | 1.55E-01 | 5.00E-02 | 4.40E-01 | 1.55E-01 | 1.30E-01 | 7.00E-02 | 0.82 | 1.46 | 1.24 | 0.82 | 0.95 | 0.71 |
| | | | | | Baseline | PBMC | Normal | 3.03E-08 | 4.71E-09 | 3.57E-08 | 1.82E-08 | 1.28E-08 | 1.01E-08 | 2.71E-01 | 4.22E-02 | 3.19E-01 | 1.63E-01 | 1.14E-01 | 9.04E-02 | | | | | | |
| | | | | | End of Treatment | PBMC | Normal | 2.19E-08 | 4.74E-09 | 3.49E-08 | 1.30E-08 | 1.60E-08 | 7.10E-09 | 2.24E-01 | 4.85E-02 | 3.58E-01 | 1.33E-01 | 1.64E-01 | 7.27E-02 | 0.83 | 1.15 | 1.12 | 0.82 | 1.43 | 0.80 |
| PMC-09 | Multiple Myeloma | Mutant (Cys238Tyr) | 100mg/m2 | 1 | Baseline | MACS Sorted Bone Marrow Aspirate: | Tumour | 7.18E-08 | 8.17E-08 | 1.17E-07 | 5.67E-08 | 7.64E-08 | 5.14E-08 | 1.58E-01 | 1.79E-01 | 2.57E-01 | 1.25E-01 | 1.68E-01 | 1.13E-01 | | | | | | |
| | | | | | End of Treatment | Tumour Cells | Tumour | 6.13E-08 | 6.68E-08 | 1.13E-07 | 5.12E-08 | 6.86E-08 | 4.67E-08 | 1.51E-01 | 1.64E-01 | 2.76E-01 | 1.26E-01 | 1.69E-01 | 1.15E-01 | 0.95 | 0.91 | 1.07 | 1.01 | 1.00 | 1.01 |
| | | | | | Baseline | MACS Sorted Bone Marrow Aspirate: | Normal | 4.77E-08 | 3.66E-08 | 1.08E-07 | 4.35E-08 | 4.94E-08 | 2.81E-08 | 1.52E-01 | 1.17E-01 | 3.45E-01 | 1.39E-01 | 1.58E-01 | 8.97E-02 | | | | | | |
| | | | | | End of Treatment | Normal Cells | Normal | 4.42E-08 | 2.49E-08 | 1.13E-07 | 4.14E-08 | 4.42E-08 | 1.80E-08 | 1.55E-01 | 8.72E-02 | 3.95E-01 | 1.45E-01 | 1.55E-01 | 6.30E-02 | 1.02 | 0.75 | 1.14 | 1.05 | 0.98 | 0.70 |
| | | | | | Baseline | PBMC | Normal | 3.51E-08 | 1.67E-08 | 9.56E-08 | 3.35E-08 | 4.14E-08 | 2.47E-08 | 1.42E-01 | 6.77E-02 | 3.87E-01 | 1.35E-01 | 1.68E-01 | 1.00E-01 | | | | | | |
| | | | | | End of Treatment | PBMC | Normal | 3.78E-08 | 3.41E-08 | 9.59E-08 | 5.35E-08 | 6.00E-08 | 3.97E-08 | 1.18E-01 | 1.06E-01 | 2.99E-01 | 1.67E-01 | 1.87E-01 | 1.24E-01 | 0.83 | 1.57 | 0.77 | 1.23 | 1.11 | 1.24 |
| PMC-14 | Multiple Myeloma | Wildtype | 170mg/m2 | 4 | Baseline | Bone Marrow Trephine | Tumour | 3.19E-08 | 2.51E-08 | 7.06E-08 | 2.66E-08 | 2.66E-08 | 2.20E-08 | 1.57E-01 | 1.24E-01 | 3.48E-01 | 1.31E-01 | 1.31E-01 | 1.09E-01 | | | | | | |
| | | | | | Cycle 1 Day 2 (+24hr post) | | Tumour | 3.55E-08 | 2.75E-08 | 8.08E-08 | 2.67E-08 | 3.39E-08 | 2.67E-08 | 1.54E-01 | 1.19E-01 | 3.50E-01 | 1.15E-01 | 1.47E-01 | 1.15E-01 | 0.98 | 0.96 | 1.00 | 0.88 | 1.12 | 1.06 |
| | | | | | End of Treatment | | Tumour | 3.07E-08 | 1.78E-08 | 5.70E-08 | 2.35E-08 | 2.42E-08 | 1.35E-08 | 1.84E-01 | 1.07E-01 | 3.42E-01 | 1.41E-01 | 1.45E-01 | 8.12E-02 | 1.17 | 0.86 | 0.98 | 1.08 | 1.11 | 0.75 |
| | | | | | Baseline | PBMC | Normal | 4.83E-08 | 2.68E-08 | 1.16E-07 | 4.12E-08 | 5.37E-08 | 2.33E-08 | 1.56E-01 | 8.69E-02 | 3.75E-01 | 1.33E-01 | 1.74E-01 | 7.53E-02 | | | | | | |
| | | | | | End of Treatment | PBMC | Normal | 5.52E-08 | 3.54E-08 | 1.24E-07 | 5.02E-08 | 5.09E-08 | 2.90E-08 | 1.60E-01 | 1.03E-01 | 3.59E-01 | 1.46E-01 | 1.48E-01 | 8.42E-02 | 1.02 | 1.18 | 0.96 | 1.09 | 0.85 | 1.12 |

**CX-5461 Cohort - Part 2** IV infusion Day 1 and Day 8 of each 28 day cycle

| | | | | | | | | SBS Mutation Frequencies | | | | | | SBS Proportions | | | | | | Ratio of SBS Proportions - Post-Treatment:Pre-Treatment | | | | | |
|---|---|---|---|---|---|---|---|---|---|---|---|---|---|---|---|---|---|---|---|---|---|---|---|---|---|
| **Patient** | **Disease** | **TP53 Status** | **Dose** | **Cycles** | **Timepoint** | **Sample ID** | **Tissue** | **C>A** | **C>G** | **C>T** | **T>A** | **T>C** | **T>G** | **C>A** | **C>G** | **C>T** | **T>A** | **T>C** | **T>G** | **C>A** | **C>G** | **C>T** | **T>A** | **T>C** | **T>G** |
| PMC-01 | Cutaneous T-cell lymphoma | N/A* | 70mg/m2 | 2 | Baseline | Skin Lesion | Tumour | 2.15E-08 | 1.63E-08 | 1.51E-07 | 2.00E-08 | 3.70E-08 | 9.63E-09 | 8.41E-02 | 6.38E-02 | 5.91E-01 | 7.83E-02 | 1.45E-01 | 3.77E-02 | | | | | | |
| | | | | | Cycle 1 Day 9 (+24hr post) | | Tumour | 1.84E-08 | 7.77E-09 | 7.49E-08 | 1.98E-08 | 2.68E-08 | 1.20E-08 | 1.15E-01 | 4.87E-02 | 4.69E-01 | 1.24E-01 | 1.68E-01 | 7.52E-02 | 1.37 | 0.76 | 0.79 | 1.58 | 1.16 | 2.00 |
| | | | | | Baseline | Normal Skin Punch | Normal | 1.76E-08 | 1.50E-08 | 2.01E-07 | 3.07E-08 | 2.68E-08 | 1.31E-08 | 5.79E-02 | 4.94E-02 | 6.61E-01 | 1.01E-01 | 8.80E-02 | 4.29E-02 | | | | | | |
| | | | | | Cycle 1 Day 9 (+24hr post) | | Normal | 3.11E-08 | 1.46E-08 | 9.24E-08 | 2.62E-08 | 3.89E-08 | 1.36E-08 | 1.43E-01 | 6.73E-02 | 4.26E-01 | 1.21E-01 | 1.79E-01 | 6.28E-02 | 2.48 | 1.36 | 0.64 | 1.20 | 2.04 | 1.46 |
| | | | | | Baseline | PBMC | Normal | 5.26E-08 | 2.75E-08 | 1.89E-07 | 4.32E-08 | 5.89E-08 | 2.90E-08 | 1.31E-01 | 6.86E-02 | 4.73E-01 | 1.08E-01 | 1.47E-01 | 7.25E-02 | | | | | | |
| | | | | | End of Treatment | PBMC | Normal | 5.31E-08 | 2.88E-08 | 1.55E-07 | 4.10E-08 | 6.79E-08 | 3.07E-08 | 1.41E-01 | 7.65E-02 | 4.12E-01 | 1.09E-01 | 1.80E-01 | 8.16E-02 | 1.07 | 1.12 | 0.87 | 1.01 | 1.23 | 1.13 |

**N/A*: insufficent DNA available to perform All haem Twist assay to determined TP53 status**

**Supplementary Table 3. Somatic double base substitution (DBS) frequency and proportion across tissue types and treatment timepoints in patients receiving CX-5461.**

**CX-5461 Cohort - Part 1** IV infusion Day 1 of each 21 day cycle

| | | | | | | | | DBS Mutation Frequencies | | | | | | | | | | DBS Proportions | | | | | | | | | | Ratio of DBS Proportions - Post-Treatment:Pre-Treatment | | | | | | | | | |
|---|---|---|---|---|---|---|---|---|---|---|---|---|---|---|---|---|---|---|---|---|---|---|---|---|---|---|---|---|---|---|---|---|---|---|---|---|---|
| Patient | Disease | TP53 Status | Dose | Cycles | Timepoint | Sample ID | Tissue | AC>NN | AT>NN | CC>NN | CG>NN | CT>NN | GC>NN | TA>NN | TC>NN | TG>NN | TT>NN | AC>NN | AT>NN | CC>NN | CG>NN | CT>NN | GC>NN | TA>NN | TC>NN | TG>NN | TT>NN | AC>NN | AT>NN | CC>NN | CG>NN | CT>NN | GC>NN | TA>NN | TC>NN | TG>NN | TT>NN |
| PMC-07 | Chronic Lymphocytic Leukaemia with Richter's transformation | Mutant (Arg248Trp) | 50mg/m2 | 1 | Baseline | MACS Sorted Bone Marrow Aspirate: Tumour Cells | Tumour | 0.00E+00 | 0.00E+00 | 9.31E-10 | 0.00E+00 | 0.00E+00 | 0.00E+00 | 0.00E+00 | 0.00E+00 | 9.31E-10 | 0.00E+00 | 0.00E+00 | 0.00E+00 | 5.00E-01 | 0.00E+00 | 0.00E+00 | 0.00E+00 | 0.00E+00 | 0.00E+00 | 5.00E-01 | 0.00E+00 | | | | | | | | | | |
| | | | | | Cycle 1 Day 2 (+24hr post) | | Tumour | 0.00E+00 | 0.00E+00 | 0.00E+00 | 0.00E+00 | 0.00E+00 | 0.00E+00 | 0.00E+00 | 0.00E+00 | 7.64E-10 | 0.00E+00 | 0.00E+00 | 0.00E+00 | 0.00E+00 | 0.00E+00 | 0.00E+00 | 0.00E+00 | 0.00E+00 | 0.00E+00 | 1.00E+00 | 0.00E+00 | NA | NA | 0.00 | NA | NA | NA | NA | NA | 2.00 | NA |
| | | | | | Baseline | MACS Sorted Bone Marrow Aspirate: Normal Cells | Normal | 0.00E+00 | 0.00E+00 | 0.00E+00 | 9.25E-10 | 0.00E+00 | 0.00E+00 | 0.00E+00 | 0.00E+00 | 9.25E-10 | 0.00E+00 | 0.00E+00 | 0.00E+00 | 0.00E+00 | 5.00E-01 | 0.00E+00 | 0.00E+00 | 0.00E+00 | 0.00E+00 | 5.00E-01 | 0.00E+00 | | | | | | | | | | |
| | | | | | Cycle 1 Day 1 (+4hr post) | | Normal | 0.00E+00 | 0.00E+00 | 0.00E+00 | 0.00E+00 | 0.00E+00 | 8.24E-10 | 0.00E+00 | 0.00E+00 | 0.00E+00 | 0.00E+00 | 0.00E+00 | 0.00E+00 | 0.00E+00 | 0.00E+00 | 0.00E+00 | 1.00E+00 | 0.00E+00 | 0.00E+00 | 0.00E+00 | 0.00E+00 | NA | NA | NA | 0.00 | NA | inf | NA | NA | 0.00 | NA |
| | | | | | Baseline | PBMC | Normal | 6.73E-10 | 6.73E-10 | 0.00E+00 | 0.00E+00 | 0.00E+00 | 6.73E-10 | 0.00E+00 | 0.00E+00 | 0.00E+00 | 0.00E+00 | 3.33E-01 | 3.33E-01 | 0.00E+00 | 0.00E+00 | 0.00E+00 | 3.33E-01 | 0.00E+00 | 0.00E+00 | 0.00E+00 | 0.00E+00 | | | | | | | | | | |
| | | | | | End of Treatment | PBMC | Normal | 0.00E+00 | 0.00E+00 | 5.92E-10 | 5.92E-10 | 0.00E+00 | 0.00E+00 | 0.00E+00 | 0.00E+00 | 0.00E+00 | 0.00E+00 | 0.00E+00 | 0.00E+00 | 5.00E-01 | 5.00E-01 | 0.00E+00 | 0.00E+00 | 0.00E+00 | 0.00E+00 | 0.00E+00 | 0.00E+00 | 0.00 | 0.00 | inf | inf | NA | 0.00 | NA | NA | NA | NA |
| PMC-09 | Multiple Myeloma | Mutant (Cys238Tyr) | 100mg/m2 | 1 | Baseline | MACS Sorted Bone Marrow Aspirate: Tumour Cells | Tumour | 1.51E-09 | 0.00E+00 | 1.51E-09 | 7.56E-10 | 7.56E-10 | 7.56E-10 | 7.56E-10 | 0.00E+00 | 7.56E-10 | 0.00E+00 | 2.22E-01 | 0.00E+00 | 2.22E-01 | 1.11E-01 | 1.11E-01 | 1.11E-01 | 1.11E-01 | 0.00E+00 | 1.11E-01 | 0.00E+00 | | | | | | | | | | |
| | | | | | End of Treatment | | Tumour | 1.83E-09 | 0.00E+00 | 5.49E-09 | 0.00E+00 | 9.15E-10 | 0.00E+00 | 0.00E+00 | 1.83E-09 | 9.15E-10 | 0.00E+00 | 1.67E-01 | 0.00E+00 | 5.00E-01 | 0.00E+00 | 8.33E-02 | 0.00E+00 | 0.00E+00 | 1.67E-01 | 8.33E-02 | 0.00E+00 | 0.75 | NA | 2.25 | 0.00 | 0.75 | 0.00 | 0.00 | inf | 0.75 | NA |
| | | | | | Baseline | MACS Sorted Bone Marrow Aspirate: Normal Cells | Normal | 0.00E+00 | 0.00E+00 | 1.70E-09 | 0.00E+00 | 1.70E-09 | 0.00E+00 | 0.00E+00 | 8.52E-10 | 0.00E+00 | 0.00E+00 | 0.00E+00 | 0.00E+00 | 4.00E-01 | 0.00E+00 | 4.00E-01 | 0.00E+00 | 0.00E+00 | 2.00E-01 | 0.00E+00 | 0.00E+00 | | | | | | | | | | |
| | | | | | End of Treatment | | Normal | 1.38E-09 | 0.00E+00 | 0.00E+00 | 1.38E-09 | 6.91E-10 | 0.00E+00 | 0.00E+00 | 0.00E+00 | 6.91E-10 | 0.00E+00 | 3.33E-01 | 0.00E+00 | 0.00E+00 | 3.33E-01 | 1.67E-01 | 0.00E+00 | 0.00E+00 | 0.00E+00 | 1.67E-01 | 0.00E+00 | inf | NA | 0.00 | inf | 0.42 | NA | NA | 0.00 | inf | NA |
| | | | | | Baseline | PBMC | Normal | 0.00E+00 | 0.00E+00 | 1.59E-09 | 0.00E+00 | 0.00E+00 | 7.97E-10 | 0.00E+00 | 7.97E-10 | 0.00E+00 | 0.00E+00 | 0.00E+00 | 0.00E+00 | 5.00E-01 | 0.00E+00 | 0.00E+00 | 2.50E-01 | 0.00E+00 | 2.50E-01 | 0.00E+00 | 0.00E+00 | | | | | | | | | | |
| | | | | | End of Treatment | PBMC | Normal | 0.00E+00 | 9.22E-10 | 9.22E-10 | 0.00E+00 | 0.00E+00 | 0.00E+00 | 0.00E+00 | 9.22E-10 | 0.00E+00 | 0.00E+00 | 0.00E+00 | 3.33E-01 | 3.33E-01 | 0.00E+00 | 0.00E+00 | 0.00E+00 | 0.00E+00 | 3.33E-01 | 0.00E+00 | 0.00E+00 | NA | inf | 0.67 | NA | NA | 0.00 | NA | 1.33 | NA | NA |
| PMC-14 | Multiple Myeloma | Wildtype | 170mg/m2 | 4 | Baseline | Bone Marrow Trephine | Tumour | 0.00E+00 | 0.00E+00 | 7.59E-10 | 0.00E+00 | 7.59E-10 | 0.00E+00 | 0.00E+00 | 1.52E-09 | 0.00E+00 | 0.00E+00 | 0.00E+00 | 0.00E+00 | 2.50E-01 | 0.00E+00 | 2.50E-01 | 0.00E+00 | 0.00E+00 | 5.00E-01 | 0.00E+00 | 0.00E+00 | | | | | | | | | | |
| | | | | | Cycle 1 Day 2 (+24hr post) | | Tumour | 8.08E-10 | 0.00E+00 | 0.00E+00 | 0.00E+00 | 0.00E+00 | 0.00E+00 | 0.00E+00 | 8.08E-10 | 8.08E-10 | 0.00E+00 | 3.33E-01 | 0.00E+00 | 0.00E+00 | 0.00E+00 | 0.00E+00 | 0.00E+00 | 0.00E+00 | 3.33E-01 | 3.33E-01 | 0.00E+00 | inf | NA | 0.00 | NA | 0.00 | NA | NA | 0.67 | inf | NA |
| | | | | | End of Treatment | | Tumour | 0.00E+00 | 0.00E+00 | 7.13E-10 | 0.00E+00 | 7.13E-10 | 7.13E-10 | 0.00E+00 | 0.00E+00 | 0.00E+00 | 7.13E-10 | 0.00E+00 | 0.00E+00 | 2.50E-01 | 0.00E+00 | 2.50E-01 | 2.50E-01 | 0.00E+00 | 0.00E+00 | 0.00E+00 | 2.50E-01 | NA | NA | 1.00 | NA | 1.00 | inf | NA | 0.00 | NA | inf |
| | | | | | Baseline | PBMC | Normal | 5.96E-10 | 0.00E+00 | 0.00E+00 | 1.19E-09 | 1.19E-09 | 0.00E+00 | 0.00E+00 | 0.00E+00 | 5.96E-10 | 5.96E-10 | 1.43E-01 | 0.00E+00 | 0.00E+00 | 2.86E-01 | 2.86E-01 | 0.00E+00 | 0.00E+00 | 0.00E+00 | 1.43E-01 | 1.43E-01 | | | | | | | | | | |
| | | | | | End of Treatment | PBMC | Normal | 0.00E+00 | 7.07E-10 | 2.12E-09 | 1.41E-09 | 1.41E-09 | 7.07E-10 | 0.00E+00 | 0.00E+00 | 2.83E-09 | 0.00E+00 | 0.00E+00 | 7.69E-02 | 2.31E-01 | 1.54E-01 | 1.54E-01 | 7.69E-02 | 0.00E+00 | 0.00E+00 | 3.08E-01 | 0.00E+00 | 0.00 | inf | inf | 0.54 | 0.54 | inf | NA | NA | 2.15 | 0.00 |

| | |
|---|---|
| **NA** | 50 |
| **inf** | 19 |
| **≥2-fold increases** | 5 |

**CX-5461 Cohort - Part 2** IV infusion Day 1 and Day 8 of each 28 day cycle

| | | | | | | | | DBS Mutation Frequencies | | | | | | | | | | DBS Proportions | | | | | | | | | | Ratio of DBS Proportions - Post-Treatment:Pre-Treatment | | | | | | | | | |
|---|---|---|---|---|---|---|---|---|---|---|---|---|---|---|---|---|---|---|---|---|---|---|---|---|---|---|---|---|---|---|---|---|---|---|---|---|---|
| Patient | Disease | TP53 Status | Dose | Cycles | Timepoint | Sample ID | Tissue | AC>NN | AT>NN | CC>NN | CG>NN | CT>NN | GC>NN | TA>NN | TC>NN | TG>NN | TT>NN | AC>NN | AT>NN | CC>NN | CG>NN | CT>NN | GC>NN | TA>NN | TC>NN | TG>NN | TT>NN | AC>NN | AT>NN | CC>NN | CG>NN | CT>NN | GC>NN | TA>NN | TC>NN | TG>NN | TT>NN |
| PMC-01 | Cutaneous T-cell lymphoma | N/A* | 70mg/m2 | 2 | Baseline | Skin Lesion | Tumour | 7.41E-10 | 0.00E+00 | 2.00E-08 | 1.48E-09 | 7.41E-10 | 0.00E+00 | 0.00E+00 | 7.41E-10 | 7.41E-10 | 0.00E+00 | 3.03E-02 | 0.00E+00 | 8.18E-01 | 6.06E-02 | 3.03E-02 | 0.00E+00 | 0.00E+00 | 3.03E-02 | 3.03E-02 | 0.00E+00 | | | | | | | | | | |
| | | | | | Cycle 1 Day 9 (+24hr post) | | Tumour | 0.00E+00 | 0.00E+00 | 4.95E-09 | 0.00E+00 | 0.00E+00 | 0.00E+00 | 7.06E-10 | 7.06E-10 | 0.00E+00 | 0.00E+00 | 0.00E+00 | 0.00E+00 | 7.78E-01 | 0.00E+00 | 0.00E+00 | 0.00E+00 | 1.11E-01 | 1.11E-01 | 0.00E+00 | 0.00E+00 | 0.00 | NA | 0.95 | 0.00 | 0.00 | NA | inf | 3.67 | 0.00 | NA |
| | | | | | Baseline | Normal Skin Punch | Normal | 6.53E-10 | 6.53E-10 | 4.18E-08 | 0.00E+00 | 1.31E-09 | 0.00E+00 | 0.00E+00 | 1.96E-09 | 0.00E+00 | 6.53E-10 | 1.39E-02 | 1.39E-02 | 8.89E-01 | 0.00E+00 | 2.78E-02 | 0.00E+00 | 0.00E+00 | 4.17E-02 | 0.00E+00 | 1.39E-02 | | | | | | | | | | |
| | | | | | Cycle 1 Day 9 (+24hr post) | | Normal | 9.72E-10 | 0.00E+00 | 9.72E-09 | 0.00E+00 | 0.00E+00 | 0.00E+00 | 0.00E+00 | 9.72E-10 | 9.72E-10 | 0.00E+00 | 7.69E-02 | 0.00E+00 | 7.69E-01 | 0.00E+00 | 0.00E+00 | 0.00E+00 | 0.00E+00 | 7.69E-02 | 7.69E-02 | 0.00E+00 | 5.54 | 0.00 | 0.87 | NA | 0.00 | NA | NA | 1.85 | inf | 0.00 |
| | | | | | Baseline | PBMC | Normal | 7.85E-10 | 0.00E+00 | 7.85E-10 | 3.93E-09 | 0.00E+00 | 2.36E-09 | 1.57E-09 | 2.36E-09 | 2.36E-09 | 7.85E-10 | 5.26E-02 | 0.00E+00 | 5.26E-02 | 2.63E-01 | 0.00E+00 | 1.58E-01 | 1.05E-01 | 1.58E-01 | 1.58E-01 | 5.26E-02 | | | | | | | | | | |
| | | | | | End of Treatment | PBMC | Normal | 0.00E+00 | 6.40E-10 | 6.40E-10 | 3.84E-09 | 6.40E-10 | 1.28E-09 | 0.00E+00 | 6.40E-10 | 6.40E-10 | 0.00E+00 | 0.00E+00 | 7.69E-02 | 7.69E-02 | 4.62E-01 | 7.69E-02 | 1.54E-01 | 0.00E+00 | 7.69E-02 | 7.69E-02 | 0.00E+00 | 0.00 | inf | 1.46 | 1.75 | inf | 0.97 | 0.00 | 0.49 | 0.49 | 0.00 |

**N/A*: insufficent DNA available to perform All haem Twist assay to determined TP53 status**

**inf: The proportion was 0 in the pre-treatment sample but non-zero post-treatment, indicating appearance after treatment.**
**NA: Both pre- and post-treatment values were zero, suggesting no occurrence of that mutation type.**

Supplementary Table 4. Somatic small insertions and deletions (INDELS) frequency and proportion across tissue types and treatment timepoints in patients receiving CX-5461.

CX-5461 Cohort - Part 1 IV infusion Day 1 of each 21 day cycle

INDEL Mutation Frequencies

| Patient | Disease | TP53 Status | Dose | Cycles | Timepoint | Sample ID | Tissue | 2bp_deletion | 2bp_deletion_with_microhomology | 2bp_insertion | 3bp_deletion | 3bp_deletion_with_microhomology | 3bp_insertion | 4bp_deletion | 4bp_deletion_with_microhomology | 4bp_insertion | 5+bp_deletion | 5+bp_deletion_with_microhomology | 5+bp_insertion | C_deletion | C_insertion | T_deletion | T_insertion |
|---|---|---|---|---|---|---|---|---|---|---|---|---|---|---|---|---|---|---|---|---|---|---|---|
| PMC-07 | Chronic Lymphocytic Leukaemia with Richter's transformation | Mutant (Arg248Trp) | 50mg/m2 | 1 | Baseline | MACS Sorted Bone Marrow Aspirate: Tumour Cells | Tumour | 1.86E-09 | 0.00E+00 | 0.00E+00 | 9.31E-10 | 0.00E+00 | 0.00E+00 | 0.00E+00 | 0.00E+00 | 0.00E+00 | 1.86E-09 | 7.45E-09 | 9.31E-09 | 1.86E-09 | 0.00E+00 | 9.31E-10 | 0.00E+00 |
| | | | | | Cycle 1 Day 2 (+24hr post) | | Tumour | 0.00E+00 | 0.00E+00 | 0.00E+00 | 0.00E+00 | 0.00E+00 | 0.00E+00 | 7.64E-10 | 0.00E+00 | 0.00E+00 | 5.35E-09 | 3.82E-09 | 8.40E-09 | 2.29E-09 | 0.00E+00 | 3.06E-09 | 0.00E+00 |
| | | | | | Baseline | MACS Sorted Bone Marrow Aspirate: Normal Cells | Normal | 9.25E-10 | 0.00E+00 | 9.25E-10 | 9.25E-10 | 0.00E+00 | 0.00E+00 | 0.00E+00 | 0.00E+00 | 0.00E+00 | 1.85E-09 | 3.70E-09 | 8.32E-09 | 1.85E-09 | 0.00E+00 | 8.32E-09 | 0.00E+00 |
| | | | | | Cycle 1 Day 1 (+4hr post) | | Normal | 0.00E+00 | 0.00E+00 | 8.24E-10 | 0.00E+00 | 0.00E+00 | 0.00E+00 | 1.65E-09 | 0.00E+00 | 0.00E+00 | 1.65E-09 | 3.29E-09 | 8.24E-09 | 1.65E-09 | 0.00E+00 | 5.76E-09 | 8.24E-10 |
| | | | | | Baseline | PBMC | Normal | 6.73E-10 | 0.00E+00 | 6.73E-10 | 1.35E-09 | 6.73E-10 | 0.00E+00 | 0.00E+00 | 0.00E+00 | 0.00E+00 | 4.04E-09 | 3.37E-09 | 1.08E-08 | 3.37E-09 | 0.00E+00 | 4.71E-09 | 6.73E-10 |
| | | | | | End of Treatment | PBMC | Normal | 1.18E-09 | 0.00E+00 | 0.00E+00 | 0.00E+00 | 5.92E-10 | 0.00E+00 | 0.00E+00 | 0.00E+00 | 0.00E+00 | 2.96E-09 | 2.37E-09 | 7.70E-09 | 2.96E-09 | 0.00E+00 | 1.78E-09 | 0.00E+00 |
| PMC-09 | Multiple Myeloma | Mutant (Cys238Tyr) | 100mg/m2 | 1 | Baseline | MACS Sorted Bone Marrow Aspirate: Tumour Cells | Tumour | 2.27E-09 | 0.00E+00 | 7.56E-10 | 0.00E+00 | 0.00E+00 | 0.00E+00 | 0.00E+00 | 0.00E+00 | 0.00E+00 | 3.78E-09 | 3.02E-09 | 9.83E-09 | 3.02E-09 | 0.00E+00 | 3.78E-09 | 3.02E-09 |
| | | | | | End of Treatment | | Tumour | 0.00E+00 | 0.00E+00 | 9.15E-10 | 0.00E+00 | 0.00E+00 | 0.00E+00 | 0.00E+00 | 0.00E+00 | 0.00E+00 | 5.49E-09 | 5.49E-09 | 7.32E-09 | 9.15E-10 | 0.00E+00 | 2.74E-09 | 9.15E-10 |
| | | | | | Baseline | MACS Sorted Bone Marrow Aspirate: Normal Cells | Normal | 8.52E-10 | 0.00E+00 | 0.00E+00 | 0.00E+00 | 0.00E+00 | 0.00E+00 | 0.00E+00 | 0.00E+00 | 0.00E+00 | 5.11E-09 | 3.41E-09 | 5.97E-09 | 5.97E-09 | 8.52E-10 | 9.37E-09 | 0.00E+00 |
| | | | | | End of Treatment | | Normal | 6.91E-10 | 6.91E-10 | 6.91E-10 | 6.91E-10 | 0.00E+00 | 0.00E+00 | 0.00E+00 | 0.00E+00 | 0.00E+00 | 2.07E-09 | 1.38E-09 | 4.83E-09 | 3.45E-09 | 0.00E+00 | 5.53E-09 | 0.00E+00 |
| | | | | | Baseline | PBMC | Normal | 7.97E-10 | 0.00E+00 | 7.97E-10 | 0.00E+00 | 0.00E+00 | 0.00E+00 | 0.00E+00 | 0.00E+00 | 0.00E+00 | 7.97E-10 | 7.97E-10 | 9.56E-09 | 1.59E-09 | 7.97E-10 | 7.97E-09 | 0.00E+00 |
| | | | | | End of Treatment | PBMC | Normal | 2.77E-09 | 0.00E+00 | 9.22E-10 | 0.00E+00 | 9.22E-10 | 0.00E+00 | 0.00E+00 | 0.00E+00 | 0.00E+00 | 4.61E-09 | 4.61E-09 | 7.38E-09 | 2.77E-09 | 0.00E+00 | 5.53E-09 | 0.00E+00 |
| PMC-14 | Multiple Myeloma | Wildtype | 170mg/m2 | 4 | Baseline | Bone Marrow Trephine | Tumour | 1.52E-09 | 0.00E+00 | 7.59E-10 | 0.00E+00 | 0.00E+00 | 0.00E+00 | 0.00E+00 | 0.00E+00 | 0.00E+00 | 4.56E-09 | 5.31E-09 | 1.67E-08 | 3.04E-09 | 1.52E-09 | 0.00E+00 | 7.59E-10 |
| | | | | | Cycle 1 Day 2 (+24hr post) | | Tumour | 1.62E-09 | 0.00E+00 | 8.08E-10 | 0.00E+00 | 8.08E-10 | 0.00E+00 | 0.00E+00 | 8.08E-10 | 0.00E+00 | 8.88E-09 | 2.42E-09 | 1.29E-08 | 4.85E-09 | 0.00E+00 | 1.62E-09 | 0.00E+00 |
| | | | | | End of Treatment | | Tumour | 7.13E-10 | 7.13E-10 | 0.00E+00 | 7.13E-10 | 0.00E+00 | 0.00E+00 | 0.00E+00 | 0.00E+00 | 0.00E+00 | 4.28E-09 | 3.56E-09 | 1.50E-08 | 2.85E-09 | 0.00E+00 | 3.56E-09 | 7.13E-10 |
| | | | | | Baseline | PBMC | Normal | 5.96E-10 | 5.96E-10 | 5.96E-10 | 0.00E+00 | 0.00E+00 | 0.00E+00 | 0.00E+00 | 0.00E+00 | 0.00E+00 | 3.58E-09 | 2.39E-09 | 8.35E-09 | 4.17E-09 | 0.00E+00 | 2.39E-09 | 0.00E+00 |
| | | | | | End of Treatment | PBMC | Normal | 1.41E-09 | 0.00E+00 | 0.00E+00 | 0.00E+00 | 0.00E+00 | 0.00E+00 | 0.00E+00 | 2.12E-09 | 0.00E+00 | 3.54E-09 | 4.95E-09 | 1.63E-08 | 7.07E-09 | 7.07E-10 | 2.12E-09 | 7.07E-10 |

INDEL Proportions

| Patient | Timepoint | Tissue | 2bp_deletion | 2bp_deletion_with_microhomology | 2bp_insertion | 3bp_deletion | 3bp_deletion_with_microhomology | 3bp_insertion | 4bp_deletion | 4bp_deletion_with_microhomology | 4bp_insertion | 5+bp_deletion | 5+bp_deletion_with_microhomology | 5+bp_insertion | C_deletion | C_insertion | T_deletion | T_insertion |
|---|---|---|---|---|---|---|---|---|---|---|---|---|---|---|---|---|---|---|
| PMC-07 | Baseline | Tumour | 7.69E-02 | 0.00E+00 | 0.00E+00 | 3.85E-02 | 0.00E+00 | 0.00E+00 | 0.00E+00 | 0.00E+00 | 0.00E+00 | 7.69E-02 | 3.08E-01 | 3.85E-01 | 7.69E-02 | 0.00E+00 | 3.85E-02 | 0.00E+00 |
| | Cycle 1 Day 2 (+24hr post) | Tumour | 0.00E+00 | 0.00E+00 | 0.00E+00 | 0.00E+00 | 0.00E+00 | 0.00E+00 | 3.23E-02 | 0.00E+00 | 0.00E+00 | 2.26E-01 | 1.61E-01 | 3.55E-01 | 9.68E-02 | 0.00E+00 | 1.29E-01 | 0.00E+00 |
| | Baseline | Normal | 3.45E-02 | 0.00E+00 | 3.45E-02 | 3.45E-02 | 0.00E+00 | 0.00E+00 | 0.00E+00 | 0.00E+00 | 0.00E+00 | 6.90E-02 | 1.38E-01 | 3.10E-01 | 6.90E-02 | 0.00E+00 | 3.10E-01 | 0.00E+00 |
| | Cycle 1 Day 1 (+4hr post) | Normal | 0.00E+00 | 0.00E+00 | 3.45E-02 | 0.00E+00 | 0.00E+00 | 0.00E+00 | 6.90E-02 | 0.00E+00 | 0.00E+00 | 6.90E-02 | 1.38E-01 | 3.45E-01 | 6.90E-02 | 0.00E+00 | 2.41E-01 | 3.45E-02 |
| | Baseline | Normal | 2.22E-02 | 0.00E+00 | 2.22E-02 | 4.44E-02 | 2.22E-02 | 0.00E+00 | 0.00E+00 | 0.00E+00 | 0.00E+00 | 1.33E-01 | 1.11E-01 | 3.56E-01 | 1.11E-01 | 0.00E+00 | 1.56E-01 | 2.22E-02 |
| | End of Treatment | Normal | 6.06E-02 | 0.00E+00 | 0.00E+00 | 0.00E+00 | 3.03E-02 | 0.00E+00 | 0.00E+00 | 0.00E+00 | 0.00E+00 | 1.52E-01 | 1.21E-01 | 3.94E-01 | 1.52E-01 | 0.00E+00 | 9.09E-02 | 0.00E+00 |
| PMC-09 | Baseline | Tumour | 7.69E-02 | 0.00E+00 | 2.56E-02 | 0.00E+00 | 0.00E+00 | 0.00E+00 | 0.00E+00 | 0.00E+00 | 0.00E+00 | 1.28E-01 | 1.03E-01 | 3.33E-01 | 1.03E-01 | 0.00E+00 | 1.28E-01 | 1.03E-01 |
| | End of Treatment | Tumour | 0.00E+00 | 0.00E+00 | 3.85E-02 | 0.00E+00 | 0.00E+00 | 0.00E+00 | 0.00E+00 | 0.00E+00 | 0.00E+00 | 2.31E-01 | 2.31E-01 | 3.08E-01 | 3.85E-02 | 0.00E+00 | 1.15E-01 | 3.85E-02 |
| | Baseline | Normal | 2.70E-02 | 0.00E+00 | 0.00E+00 | 0.00E+00 | 0.00E+00 | 0.00E+00 | 0.00E+00 | 0.00E+00 | 0.00E+00 | 1.62E-01 | 1.08E-01 | 1.89E-01 | 1.89E-01 | 2.70E-02 | 2.97E-01 | 0.00E+00 |
| | End of Treatment | Normal | 3.45E-02 | 3.45E-02 | 3.45E-02 | 3.45E-02 | 0.00E+00 | 0.00E+00 | 0.00E+00 | 0.00E+00 | 0.00E+00 | 1.03E-01 | 6.90E-02 | 2.41E-01 | 1.72E-01 | 0.00E+00 | 2.76E-01 | 0.00E+00 |
| | Baseline | Normal | 3.45E-02 | 0.00E+00 | 3.45E-02 | 0.00E+00 | 0.00E+00 | 0.00E+00 | 0.00E+00 | 0.00E+00 | 0.00E+00 | 3.45E-02 | 3.45E-02 | 4.14E-01 | 6.90E-02 | 3.45E-02 | 3.45E-01 | 0.00E+00 |
| | End of Treatment | Normal | 9.38E-02 | 0.00E+00 | 3.13E-02 | 0.00E+00 | 3.13E-02 | 0.00E+00 | 0.00E+00 | 0.00E+00 | 0.00E+00 | 1.56E-01 | 1.56E-01 | 2.50E-01 | 9.38E-02 | 0.00E+00 | 1.88E-01 | 0.00E+00 |
| PMC-14 | Baseline | Tumour | 4.44E-02 | 0.00E+00 | 2.22E-02 | 0.00E+00 | 0.00E+00 | 0.00E+00 | 0.00E+00 | 0.00E+00 | 0.00E+00 | 1.33E-01 | 1.56E-01 | 4.89E-01 | 8.89E-02 | 4.44E-02 | 0.00E+00 | 2.22E-02 |
| | Cycle 1 Day 2 (+24hr post) | Tumour | 4.65E-02 | 0.00E+00 | 2.33E-02 | 0.00E+00 | 2.33E-02 | 0.00E+00 | 0.00E+00 | 2.33E-02 | 0.00E+00 | 2.56E-01 | 6.98E-02 | 3.72E-01 | 1.40E-01 | 0.00E+00 | 4.65E-02 | 0.00E+00 |
| | End of Treatment | Tumour | 2.22E-02 | 2.22E-02 | 0.00E+00 | 2.22E-02 | 0.00E+00 | 0.00E+00 | 0.00E+00 | 0.00E+00 | 0.00E+00 | 1.33E-01 | 1.11E-01 | 4.67E-01 | 8.89E-02 | 0.00E+00 | 1.11E-01 | 2.22E-02 |
| | Baseline | Normal | 2.63E-02 | 2.63E-02 | 2.63E-02 | 0.00E+00 | 0.00E+00 | 0.00E+00 | 0.00E+00 | 0.00E+00 | 0.00E+00 | 1.58E-01 | 1.05E-01 | 3.68E-01 | 1.84E-01 | 0.00E+00 | 1.05E-01 | 0.00E+00 |
| | End of Treatment | Normal | 3.64E-02 | 0.00E+00 | 0.00E+00 | 0.00E+00 | 0.00E+00 | 0.00E+00 | 0.00E+00 | 5.45E-02 | 0.00E+00 | 9.09E-02 | 1.27E-01 | 4.18E-01 | 1.82E-01 | 1.82E-02 | 5.45E-02 | 1.82E-02 |

Ratio of INDEL Proportions - Post-Treatment:Pre-Treatment

| Patient | Timepoint | Tissue | 2bp_deletion | 2bp_deletion_with_microhomology | 2bp_insertion | 3bp_deletion | 3bp_deletion_with_microhomology | 3bp_insertion | 4bp_deletion | 4bp_deletion_with_microhomology | 4bp_insertion | 5+bp_deletion | 5+bp_deletion_with_microhomology | 5+bp_insertion | C_deletion | C_insertion | T_deletion | T_insertion |
|---|---|---|---|---|---|---|---|---|---|---|---|---|---|---|---|---|---|---|
| PMC-07 | Baseline | Tumour | | | | | | | | | | | | | | | | |
| | Cycle 1 Day 2 (+24hr post) | Tumour | 0.00 | NA | NA | 0.00 | NA | NA | inf | NA | NA | 2.94 | 0.52 | 0.92 | 1.26 | NA | 3.35 | NA |
| | Baseline | Normal | | | | | | | | | | | | | | | | |
| | Cycle 1 Day 1 (+4hr post) | Normal | 0.00 | NA | 1.00 | 0.00 | NA | NA | inf | NA | NA | 1.00 | 1.00 | 1.11 | 1.00 | NA | 0.78 | inf |
| | Baseline | Normal | | | | | | | | | | | | | | | | |
| | End of Treatment | Normal | 2.73 | NA | 0.00 | 0.00 | 1.36 | NA | NA | NA | NA | 1.14 | 1.09 | 1.11 | 1.36 | NA | 0.58 | 0.00 |
| PMC-09 | Baseline | Tumour | | | | | | | | | | | | | | | | |
| | End of Treatment | Tumour | 0.00 | NA | 1.50 | NA | NA | NA | NA | NA | NA | 1.80 | 2.25 | 0.92 | 0.38 | NA | 0.90 | 0.38 |
| | Baseline | Normal | | | | | | | | | | | | | | | | |
| | End of Treatment | Normal | 1.28 | inf | inf | inf | NA | NA | NA | NA | NA | 0.64 | 0.64 | 1.28 | 0.91 | 0.00 | 0.93 | NA |
| | Baseline | Normal | | | | | | | | | | | | | | | | |
| | End of Treatment | Normal | 2.72 | NA | 0.91 | NA | inf | NA | NA | NA | NA | 4.53 | 4.53 | 0.60 | 1.36 | 0.00 | 0.54 | NA |
| PMC-14 | Baseline | Tumour | | | | | | | | | | | | | | | | |
| | Cycle 1 Day 2 (+24hr post) | Tumour | 1.05 | NA | 1.05 | NA | inf | NA | NA | inf | NA | 1.92 | 0.45 | 0.76 | 1.57 | 0.00 | inf | 0.00 |
| | End of Treatment | Tumour | 0.50 | inf | 0.00 | inf | NA | NA | NA | NA | NA | 1.00 | 0.71 | 0.95 | 1.00 | 0.00 | inf | 1.00 |
| | Baseline | Normal | | | | | | | | | | | | | | | | |
| | End of Treatment | Normal | 1.38 | 0.00 | 0.00 | NA | NA | NA | NA | inf | NA | 0.58 | 1.21 | 1.14 | 0.99 | inf | 0.52 | inf |

| | |
|---|---|
| NA | 68 |
| inf | 22 |
| ≥2-fold increases | 8 |

CX-5461 Cohort - Part 2 IV infusion Day 1 and Day 8 of each 28 day cycle

INDEL Mutation Frequencies

| Patient | Disease | TP53 Status | Dose | Cycles | Timepoint | Sample ID | Tissue | 2bp_deletion | 2bp_deletion_with_microhomology | 2bp_insertion | 3bp_deletion | 3bp_deletion_with_microhomology | 3bp_insertion | 4bp_deletion | 4bp_deletion_with_microhomology | 4bp_insertion | 5+bp_deletion | 5+bp_deletion_with_microhomology | 5+bp_insertion | C_deletion | C_insertion | T_deletion | T_insertion |
|---|---|---|---|---|---|---|---|---|---|---|---|---|---|---|---|---|---|---|---|---|---|---|---|
| PMC-01 | Cutaneous T-cell lymphoma | N/A* | 70mg/m2 | 2 | Baseline | Skin Lesion | Tumour | 1.48E-09 | 0.00E+00 | 0.00E+00 | 0.00E+00 | 0.00E+00 | 0.00E+00 | 0.00E+00 | 0.00E+00 | 0.00E+00 | 8.15E-09 | 4.45E-09 | 1.04E-08 | 5.19E-09 | 7.41E-10 | 5.93E-09 | 7.41E-10 |
| | | | | | Cycle 1 Day 9 (+24hr post) | | Tumour | 7.06E-10 | 0.00E+00 | 0.00E+00 | 7.06E-10 | 0.00E+00 | 0.00E+00 | 0.00E+00 | 0.00E+00 | 0.00E+00 | 7.77E-09 | 9.18E-09 | 6.36E-09 | 0.00E+00 | 7.06E-10 | 6.36E-09 | 0.00E+00 |
| | | | | | Baseline | Normal Skin Punch | Normal | 0.00E+00 | 0.00E+00 | 6.53E-10 | 0.00E+00 | 0.00E+00 | 0.00E+00 | 0.00E+00 | 6.53E-10 | 6.53E-10 | 3.92E-09 | 5.22E-09 | 9.79E-09 | 3.26E-09 | 6.53E-10 | 2.61E-09 | 1.96E-09 |
| | | | | | Cycle 1 Day 9 (+24hr post) | | Normal | 9.72E-10 | 9.72E-10 | 9.72E-10 | 9.72E-10 | 9.72E-10 | 0.00E+00 | 0.00E+00 | 0.00E+00 | 0.00E+00 | 8.75E-09 | 7.78E-09 | 1.17E-08 | 1.94E-09 | 0.00E+00 | 4.86E-09 | 9.72E-10 |
| | | | | | Baseline | PBMC | Normal | 7.85E-10 | 1.57E-09 | 7.85E-10 | 0.00E+00 | 7.85E-10 | 0.00E+00 | 7.85E-10 | 7.85E-10 | 0.00E+00 | 6.28E-09 | 3.14E-09 | 7.07E-09 | 3.14E-09 | 0.00E+00 | 5.50E-09 | 7.85E-10 |
| | | | | | End of Treatment | PBMC | Normal | 1.28E-09 | 6.40E-10 | 6.40E-10 | 6.40E-10 | 6.40E-10 | 0.00E+00 | 0.00E+00 | 6.40E-10 | 0.00E+00 | 3.20E-09 | 6.40E-09 | 7.68E-09 | 3.20E-09 | 0.00E+00 | 7.68E-09 | 1.28E-09 |

INDEL Proportions

| Patient | Timepoint | Tissue | 2bp_deletion | 2bp_deletion_with_microhomology | 2bp_insertion | 3bp_deletion | 3bp_deletion_with_microhomology | 3bp_insertion | 4bp_deletion | 4bp_deletion_with_microhomology | 4bp_insertion | 5+bp_deletion | 5+bp_deletion_with_microhomology | 5+bp_insertion | C_deletion | C_insertion | T_deletion | T_insertion |
|---|---|---|---|---|---|---|---|---|---|---|---|---|---|---|---|---|---|---|
| PMC-01 | Baseline | Tumour | 4.00E-02 | 0.00E+00 | 0.00E+00 | 0.00E+00 | 0.00E+00 | 0.00E+00 | 0.00E+00 | 0.00E+00 | 0.00E+00 | 2.20E-01 | 1.20E-01 | 2.80E-01 | 1.40E-01 | 2.00E-02 | 1.60E-01 | 2.00E-02 |
| | Cycle 1 Day 9 (+24hr post) | Tumour | 2.22E-02 | 0.00E+00 | 0.00E+00 | 2.22E-02 | 0.00E+00 | 0.00E+00 | 0.00E+00 | 0.00E+00 | 0.00E+00 | 2.44E-01 | 2.89E-01 | 2.00E-01 | 0.00E+00 | 2.22E-02 | 2.00E-01 | 0.00E+00 |
| | Baseline | Normal | 0.00E+00 | 0.00E+00 | 2.22E-02 | 0.00E+00 | 0.00E+00 | 0.00E+00 | 0.00E+00 | 2.22E-02 | 2.22E-02 | 1.33E-01 | 1.78E-01 | 3.33E-01 | 1.11E-01 | 2.22E-02 | 8.89E-02 | 6.67E-02 |
| | Cycle 1 Day 9 (+24hr post) | Normal | 2.38E-02 | 2.38E-02 | 2.38E-02 | 2.38E-02 | 2.38E-02 | 0.00E+00 | 0.00E+00 | 0.00E+00 | 0.00E+00 | 2.14E-01 | 1.90E-01 | 2.86E-01 | 4.76E-02 | 0.00E+00 | 1.19E-01 | 2.38E-02 |
| | Baseline | Normal | 2.50E-02 | 5.00E-02 | 2.50E-02 | 0.00E+00 | 2.50E-02 | 0.00E+00 | 2.50E-02 | 2.50E-02 | 0.00E+00 | 2.00E-01 | 1.00E-01 | 2.25E-01 | 1.00E-01 | 0.00E+00 | 1.75E-01 | 2.50E-02 |
| | End of Treatment | Normal | 3.77E-02 | 1.89E-02 | 1.89E-02 | 1.89E-02 | 1.89E-02 | 0.00E+00 | 0.00E+00 | 1.89E-02 | 0.00E+00 | 9.43E-02 | 1.89E-01 | 2.26E-01 | 9.43E-02 | 0.00E+00 | 2.26E-01 | 3.77E-02 |

Ratio of INDEL Proportions - Post-Treatment:Pre-Treatment

| Patient | Timepoint | Tissue | 2bp_deletion | 2bp_deletion_with_microhomology | 2bp_insertion | 3bp_deletion | 3bp_deletion_with_microhomology | 3bp_insertion | 4bp_deletion | 4bp_deletion_with_microhomology | 4bp_insertion | 5+bp_deletion | 5+bp_deletion_with_microhomology | 5+bp_insertion | C_deletion | C_insertion | T_deletion | T_insertion |
|---|---|---|---|---|---|---|---|---|---|---|---|---|---|---|---|---|---|---|
| PMC-01 | Baseline | Tumour | | | | | | | | | | | | | | | | |
| | Cycle 1 Day 9 (+24hr post) | Tumour | 0.56 | NA | NA | inf | NA | NA | NA | NA | NA | 1.11 | 2.41 | 0.71 | 0.00 | 1.11 | 1.25 | 0.00 |
| | Baseline | Normal | | | | | | | | | | | | | | | | |
| | Cycle 1 Day 9 (+24hr post) | Normal | inf | inf | 1.07 | inf | inf | NA | NA | 0.00 | 0.00 | 1.61 | 1.07 | 0.86 | 0.43 | 0.00 | 1.34 | 0.36 |
| | Baseline | Normal | | | | | | | | | | | | | | | | |
| | End of Treatment | Normal | 1.51 | 0.38 | 0.75 | inf | 0.75 | NA | 0.00 | 0.75 | NA | 0.47 | 1.89 | 1.01 | 0.94 | NA | 1.29 | 1.51 |

N/A*: insufficent DNA available to perform All haem Twist assay to determined TP53 status

inf: The proportion was 0 in the pre-treatment sample but non-zero post-treatment, indicating appearance after treatment.
NA: Both pre- and post-treatment values were zero, suggesting no occurrence of that mutation type.
